# Attention Deficit Hyperactivity Disorder Symptoms and Brain Morphology: Examining Confounding Bias

**DOI:** 10.1101/2022.05.03.22274414

**Authors:** Lorenza Dall’Aglio, Hannah H. Kim, Sander Lamballais, Jeremy Labrecque, Ryan L. Muetzel, Henning Tiemeier

**Author notes:** **Corresponding author:** Henning Tiemeier, MD Ph.D.; Department of Social and Behavioral Sciences, Harvard T.H. Chan School of Public Health, 677 Huntington Ave, Boston, 02115 MA, USA. Co-first author (Lorenza Dall’Aglio, Hannah H. Kim, Sander Lamballais). Co-last author (Ryan L. Muetzel, Henning Tiemeier).

## Abstract

**Background:** Associations between attention-deficit/hyperactivity disorder (ADHD) and brain morphology have been reported, although with several inconsistencies. These may partly stem from confounding bias, which could distort associations and limit generalizability. We examined how associations between brain morphology and ADHD symptoms change with adjustments for potential confounders typically overlooked in the literature (aim 1), and for IQ, which is typically corrected for but plays an unclear role (aim 2).

**Methods:** Participants were 10-year-old children from the Adolescent Brain Cognitive Development (*N*=7,961) and Generation R (*N*=2,531) studies. Cortical area and volume were measured with MRI and ADHD symptoms with the Child Behavior Checklist. Surface-based cross-sectional analyses were run.

**Results:** ADHD symptoms related to widespread cortical regions when solely adjusting for demographic factors. Additional adjustments for socioeconomic and maternal behavioral confounders (aim 1) generally attenuated associations, as cluster sizes halved and effect sizes substantially reduced. Cluster sizes were further reduced when including IQ (aim 2), however, we argue that adjustments could have introduced bias (e.g., by conditioning on a collider).

**Conclusions:** Careful confounder selection and control can help identify more robust and specific regions of associations for ADHD symptoms, across two cohorts. We provided guidance to minimizing confounding bias in psychiatric neuroimaging.

**Funding:** Authors are supported by an NWO-VICI grant (NWO-ZonMW: 016.VICI.170.200 to HT) for HT, LDA, SL, and the Sophia Foundation S18-20, and Erasmus University and Erasmus MC Fellowship for RLM.

## Introduction

Large strides have been made in the identification of neuroanatomical correlates of psychiatric problems, with Attention-Deficit/Hyperactivity Disorder (ADHD) being a prominent example. ADHD is the most prevalent neurodevelopmental disorder in children worldwide and is characterized by atypical levels of inattention, hyperactivity, and/or impulsivity (1). Structural Magnetic Resonance Imaging (sMRI) studies have highlighted that children with ADHD show widespread morphological differences, such as in the basal ganglia (2), subcortical areas (3), and frontal, cingulate, and temporal cortices compared to children without the disorder (4,5).

Consistently identifying the neuroanatomical substrate of ADHD, however, remains challenging. A recent meta-analysis did not find convergence across the literature on brain differences in children and adolescents with ADHD (6). One possible explanation for this inconsistency is the multifaceted nature of ADHD, in which children with the disorder have heterogeneous presentations on several cognitive and emotional domains, which could stem from distinct brain structural substrates. Other explanations regard study design. If suboptimal, it may lead to biased estimates and lack of generalizability, thus potentially concealing robust and replicable relations of brain morphology with ADHD. The present study focuses on confounding, a common source of bias in etiological studies.

Confounding bias arises when a third variable affects both the determinant (independent variable) and outcome (dependent variable) of interest (i.e., is a common cause) (7). Confounding leads to over-or under-estimation of the true effect between determinant and outcome and can even change the direction of an association. To minimize confounding bias, appropriate confounder control is paramount, although it is challenging, especially in observational studies like most neuroimaging studies of ADHD. Previous literature and expert knowledge can guide the identification of potential confounders (8), which can then be appropriately adjusted for in regression models or using methods such as restriction, standardization, or propensity scores.

Within neuroimaging studies of ADHD, except for a few large investigations (3,9,10), studies have generally matched or adjusted for a few demographic variables (e.g., age, sex) and neuroimaging metrics or parameters. Of the 19 studies included in a systematic review of neuroimaging studies on ADHD (11), 17 adjusted or matched for age in their analyses, 14 for sex, 9 for precision variables like study site, and 8 for the intelligence quotient (IQ) (**Supplementary Table 1**). Further potential confounders should, however, be considered. For instance, socioeconomic status (SES) is related to both higher risk for ADHD and variation in cortical brain structure (12,13). Thus, it is likely a confounder. Lack of adjustment for SES may have therefore concealed key relations between ADHD and brain structure. Adjustment choices are dependent on the availability of large samples with data on a wide variety of covariates, which has to date been limited for psychiatric neuroimaging studies. Yet, this is rapidly changing with the advent of population neuroscience, which entails large-scale studies with neurobiological data. This lends new opportunities for further confounder adjustments to be considered in neuroimaging studies of ADHD. Conversely, previous studies have adjusted for IQ, which may not be a confounder in the association between ADHD symptoms and the brain, and may thus have led to further bias in the results (14).

In this study, we examined the association between brain structure and ADHD symptoms and how the selection and control for potential confounders may affect results (aim 1). Moreover, we discussed the unclear role of IQ in brain structure – ADHD associations and the potential consequences of adjusting for it (aim 2). We leveraged two large, population-based cohorts: the Adolescent Brain Cognitive Development (ABCD) and the Generation R Studies. In line with most neuroimaging studies, we adopted a cross-sectional design.

## Results

### Associations between ADHD symptoms and brain morphology are widespread

We analyzed data from 10-year-old children from the ABCD (*N* = 7,961, multi-site) and Generation R (*N* = 2,531, single-site) Studies (**Supplementary Table 2**). ADHD symptoms were measured with the Child Behavioral Checklist (CBCL). T_1_-weighted images were obtained with 3T scanners (15,16). Cortical surface area and volume were considered, while thickness was not tested due to previously reported null findings in the Generation R Study (4). We ran vertex-wise linear regression models for ADHD with cortical surface area and volume. We adjusted for demographic and study characteristics which have been generally considered by previous literature (**Supplementary Table 1**): age, sex, ethnicity, and study site (ABCD only). We refer to this model as model 1, as further adjustments for confounders are outlined in subsequent steps.

We found that higher ADHD symptoms were associated with less bilateral surface area and cortical volume in both cohorts. As shown in **Figure 1**, associations were widespread for both surface area (ABCD = 1,109.8 cm^2^; GenR = 444.7 cm^2^) and volume (ABCD = 666.0 cm^2^; GenR = 96.1 cm^2^). Across both cohorts, we consistently identified clusters for surface area in the lateral occipital, postcentral, rostral middle and superior frontal, and superior parietal cortices. For volume, we observed overlap across cohorts in the cuneus, precuneus, fusiform, inferior parietal, inferior, middle, and superior temporal, isthmus of the cingulate, lateral occipital, pericalcarine, pre- and post-central, and supramarginal cortices.

**Figure 1.**
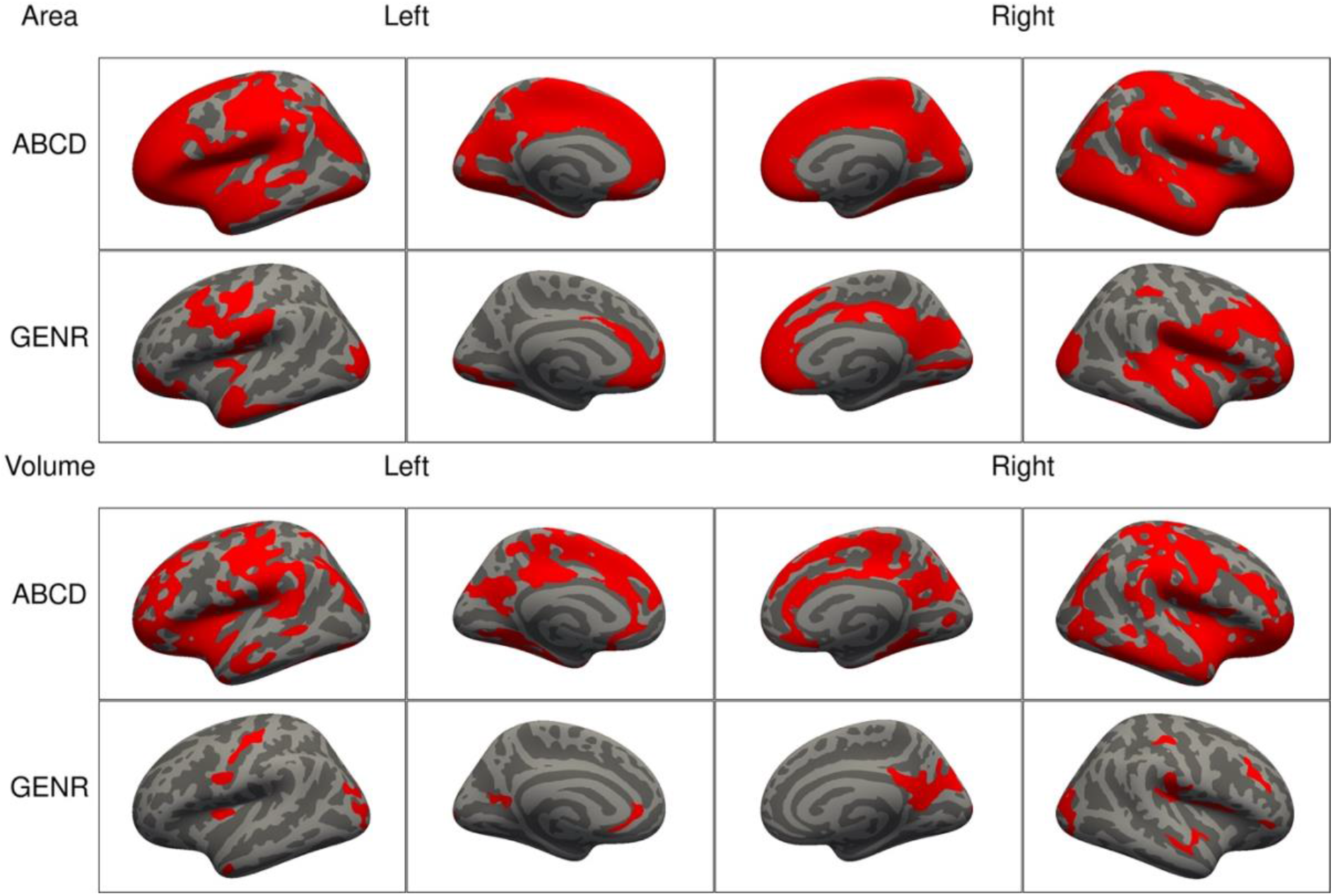
Significant clusters in the association of ADHD symptoms with cortical surface area (top) and volume (bottom) based on the ABCD and Generation R Studies, for model 1. *Note*. Rows represent the results for the ABCD or Generation R Studies, and the columns represent the left and right hemispheres. Regions in red represent significant clusters from model 1 (adjusted for sex, age, race/ethnicity).

### Confounder selection: Socioeconomic and maternal behavioral factors

Next, we considered factors that have been previously linked to ADHD and brain structure in the literature, and are thus potential confounders. To illustrate this background knowledge and the assumptions about relations between variables, we used Directed Acyclic Graphs (DAGs), a type of causal diagram (8). These guide the identification (and dismissal) of covariates that may act as confounders (**Supplementary Box 1**). Of note, while assumptions may not hold, this theoretical approach is preferred to methods selecting confounders based on model statistics (17). The DAGs are depicted in **Figure 2** and **Supplementary Figure 1**, and the rationales for variable inclusion are explained below and in the *Methods* section.

**Figure 2.**
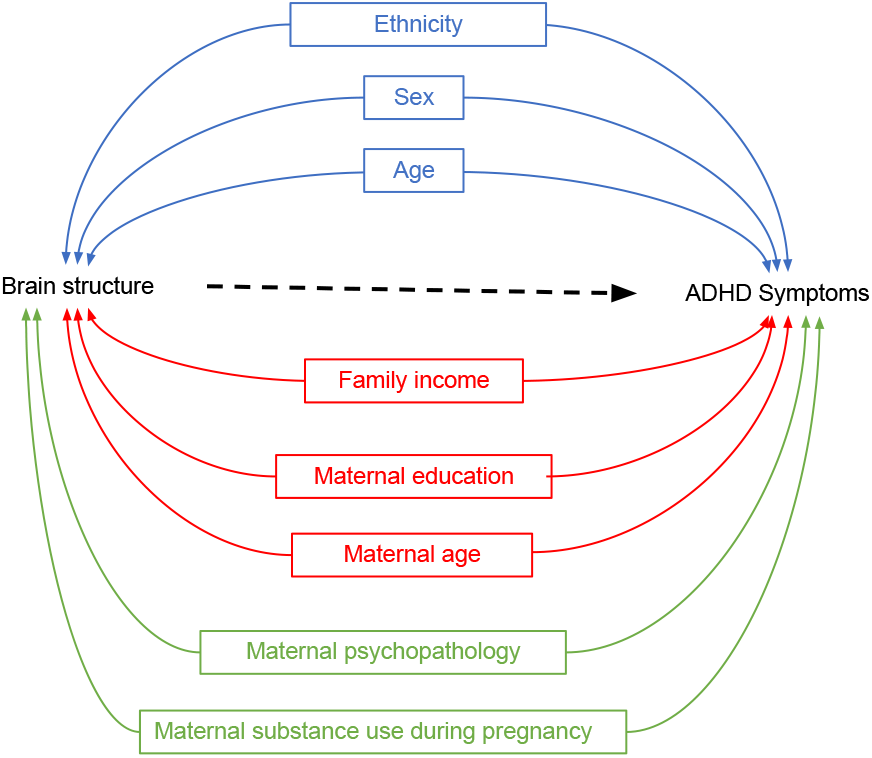
Directed Acyclic Graphs for brain structure and ADHD symptoms (simplified). *Note*. DAGs illustrating potential confounders in the association between brain structure and ADHD symptoms for three *sequential* models. Model 1 included demographic and study characteristics: Sex, age, ethnicity, study site (ABCD only) (in blue). Model 2 additionally included socioeconomic status factors: Family income, maternal education, and maternal age at childbirth (in red). Model 3 additionally incorporated postnatal maternal psychopathology and maternal substance use during pregnancy (in green).

Based on the literature, lower SES is associated with a higher risk for ADHD (12) and with variation in cortical brain structure (13). Thus, confounding by socioeconomic factors in the relation between ADHD and brain morphology is likely. We therefore additionally adjusted for a second set of confounders (model 2) related to SES: household income, maternal education, and maternal age at childbirth.

Moreover, several factors concerning maternal behavior, pre- and postnatally, have been associated with both ADHD and brain morphology. For instance, prenatal exposure to substances is known to increase the risk of developing ADHD symptoms and has been associated with variation in cerebral volume and surface area (18,19). Postnatal maternal psychopathology has been linked to higher child ADHD symptoms (20) and smaller brain volume in children (21). Thus, in model 3 we additionally adjusted for prenatal exposure to substance use (tobacco and cannabis), and postnatal maternal psychopathology.

### Adjusting for additional confounders led to reductions in the clusters of association

Adjustments for SES (model 2) led to reductions in the spatial extent of the clusters for surface area and volume in both cohorts (**Figure 3**). For surface area, cluster sizes for ADHD symptoms reduced from 1,109.8 cm^2^ in model 1 to 885.4 cm^2^ in model 2 (= - 20%) in the ABCD Study, and from 444.7 cm^2^ to 226.0 cm^2^ (= - 49%) in the Generation R Study. In the analyses of volume, clusters for ADHD symptoms were reduced from 666.0 cm^2^ in model 1 to 384.6 cm^2^ in model 2 (= - 42%) in the ABCD Study, and from 96.1 cm^2^ to 31.6 cm^2^ (= - 67%) in the Generation R Study. Adjustments for maternal substance use and psychopathology (model 3) showed further cluster changes for surface area and volume in both cohorts (**Figure 3**). For surface area, clusters covered 682.6 cm^2^ in the ABCD study (= - 23%, compared to model 2) and 214.1 cm^2^ (= - 5%) in the Generation R Study. Cortical volume clusters related to ADHD symptoms comprised 262.5 cm^2^ (= - 32%, compared to model 2) in the ABCD Study, and 33.8 cm^2^ (= + 7%) in the Generation R Study.

**Figure 3.**
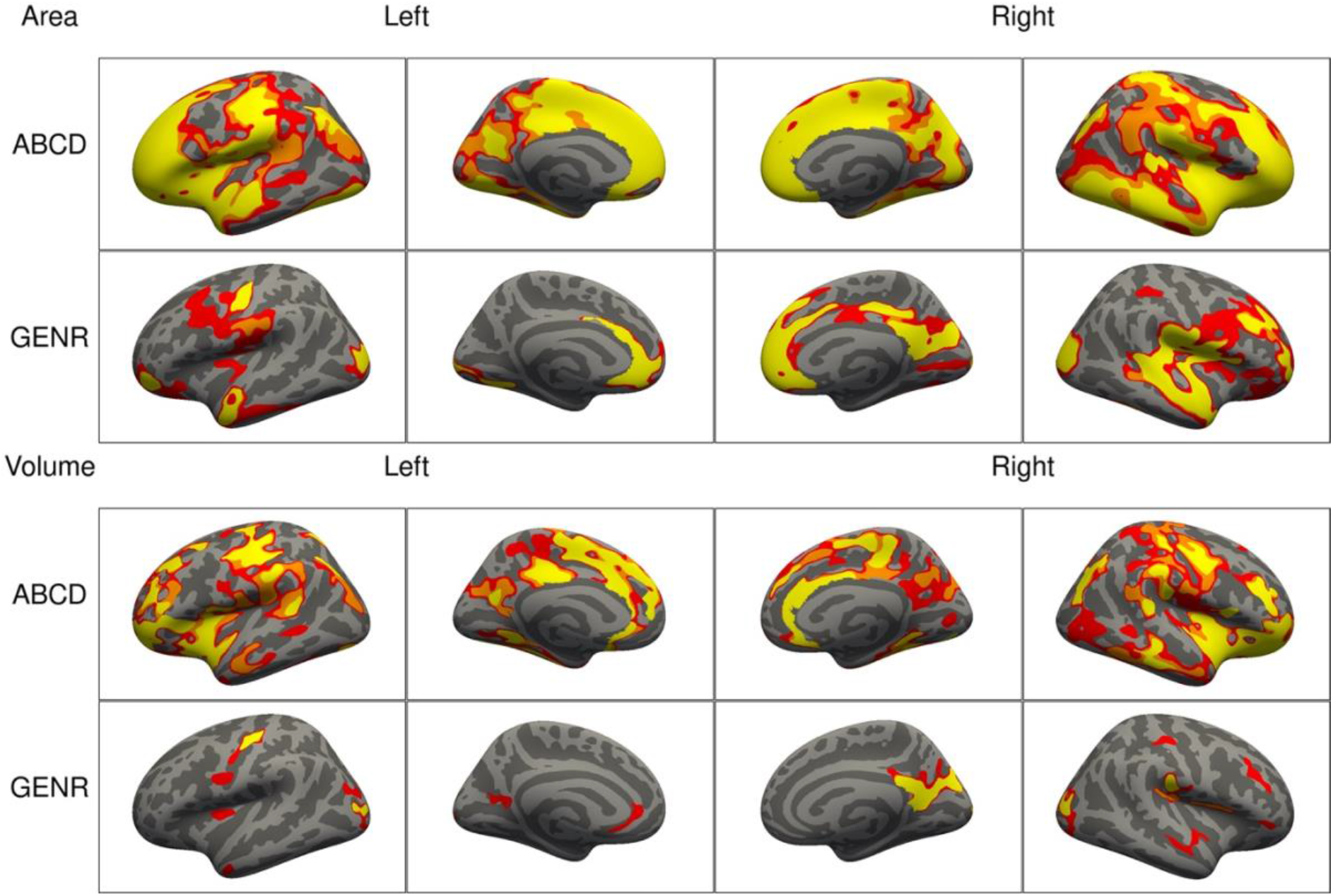
Significant clusters in the association of ADHD symptoms with cortical surface area (top) and volume (bottom) based on the ABCD and Generation R Studies, for models 1 to 3. *Note*. Rows represent the results for the ABCD or Generation R Studies, and the columns represent the left and right hemispheres. The colors denote the different models. Regions in red represent significant clusters from model 1 (sex, age, race/ethnicity), orange from model 2 (model 1 + family income, maternal education, and maternal age at childbirth), and yellow from model 3 (model 2 + maternal smoking, substance use during pregnancy, psychopathology).

After adjusting for the confounders added in model 3, across both cohorts, we consistently identified clusters for surface area in the cuneus, precuneus, fusiform, inferior parietal, isthmus of the cingulate, pericalcarine, pre- and post-central, rostral middle and superior frontal, superior temporal and supramarginal cortices. Clusters consistently identified across cohorts for volume were in the inferior parietal, isthmus of the cingulate, later occipital, pre- and post-central, precuneus, and supramarginal cortices.

### Similar results were observed for ADHD diagnosis

To explore whether the results observed for associations between brain morphology and ADHD symptoms applied to children with an ADHD diagnosis, we repeated the primary analysis using the ADHD diagnostic data from the Kiddie Schedule for Affective Disorders and Schizophrenia (KSADS) in the ABCD Study. In line with our primary results, ADHD diagnosis was associated with less bilateral surface area and volume. Compared to clusters for ADHD symptoms, those associated with ADHD diagnosis were smaller, but overlapping (**Supplementary Figure 2**). We observed similar patterns of reduction in the spatial extent of the clusters after adjusting for each set of confounders (**Supplementary Figure 3**). For surface area, cluster sizes for ADHD symptoms covered 275.5 cm^2^ in model 1 and reduced to 233.0 cm^2^ in model 2 (= - 15%), and 94.9 cm^2^ in model 3 (= - 59%, compared to model 2). For volume, cluster sizes for ADHD symptoms comprised 98.9 cm^2^ in model 1 and reduced to 70.7 cm^2^ in model 2 (= - 29%), and 26.2 cm^2^ in model 3 (= - 63%, compared to model 2).

### Beta coefficients generally decreased after confounder adjustments, but may also increase

Surface-based studies generally focus on the spatial extent of cortical clusters associated with the phenotype, but, in this study, we also explored how confounding adjustments affected the vertex-wise regression coefficients for ADHD symptoms.

At a vertex-wise level, adjusting for socioeconomic and maternal factors (model 3) led to reductions in the beta coefficients, across the brain, for both cohorts (**Supplementary Figure 4**). Of note, some beta coefficients also showed increases.

As confounding bias may lead to under-or over-estimation, it is not surprising to observe both decreases and increases in the average beta coefficients after adjustments.

At an anatomical region level, where estimates of vertices within a given Desikan-Killiany region were averaged, beta coefficients for surface area tended to decrease from model 1 to 2 by approximately 15% (**Figure 4, Supplementary Figure 5**). Further adjustments from model 2 to 3 led to decreases in the average beta coefficients of certain regions and increases in others. Similar patterns were found for volume across both studies (**Supplementary Figure 6, 7**). The average beta coefficients per region correlated moderately to strongly between the ABCD and Generation R Studies for both surface area (r_M1_ = 0.83, r_M2_ = 0.79, r_M3_ = 0.77) and volume (r_M1_ = 0.57, r_M2_ = 0.56, r_M3_ = 0.61) (**Supplementary Figure 8**).

**Figure 4.**
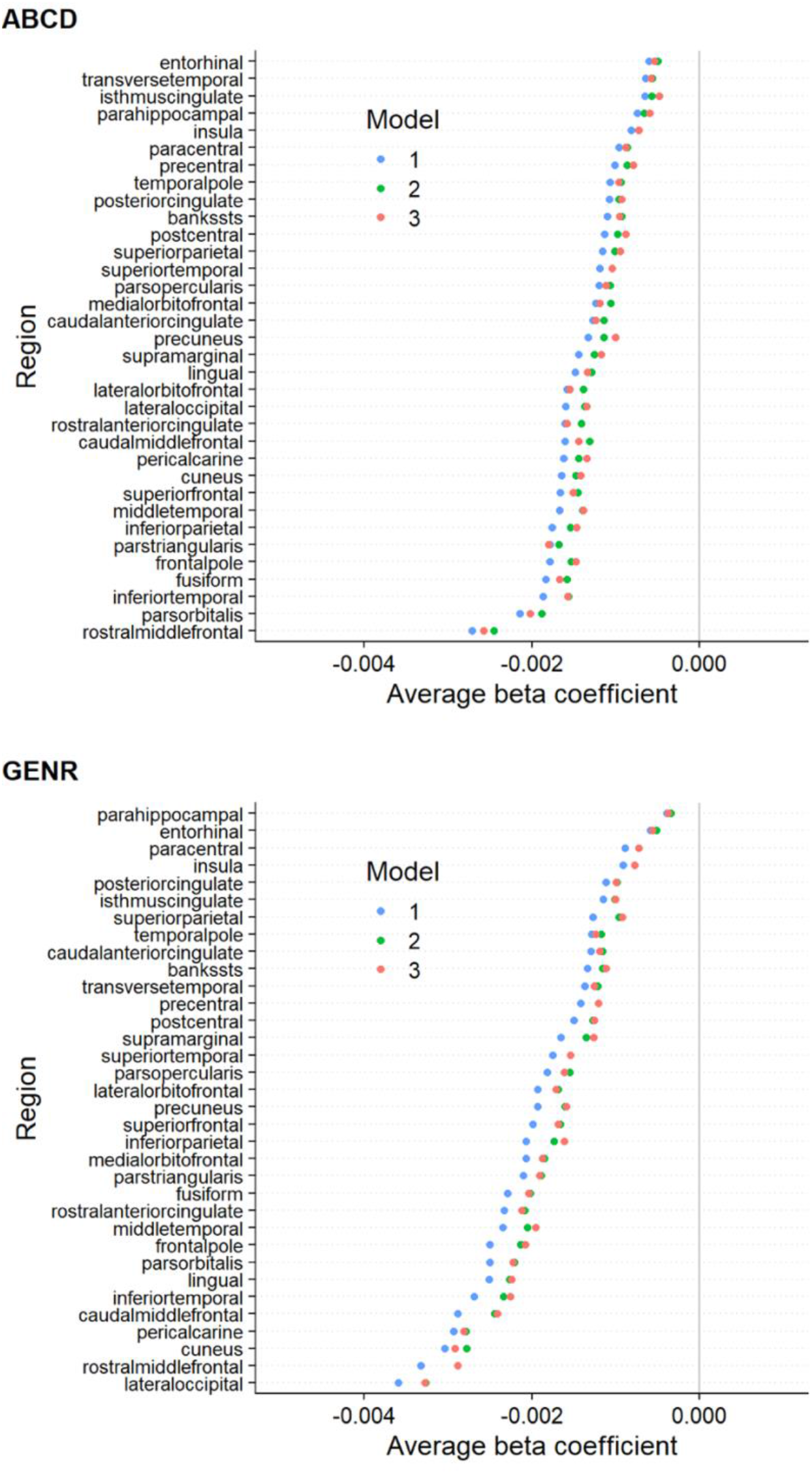
Region-based average regression coefficients for surface area in the ABCD and Generation R Studies. *Note*. The colors denote the different models, and the circles denote the average of all the betas within that region. The regions are based on the Desikan-Killiany atlas. Results for the ABCD and Generation R Studies are respectively shown on the top and bottom.

### IQ may be a confounder, mediator, or collider in neuroanatomical studies of ADHD

We considered one additional scenario which included IQ, a factor that is often adjusted for in previous studies (**Supplementary Table 1**). However, based on prior literature, it holds an ambiguous role in structural anatomy - ADHD relations. Previous studies found that children with ADHD scored lower on IQ than children without ADHD (22). Differential brain structure with levels of IQ has also been shown (23). However, the directions of causation between these variables remain unclear (24). IQ may therefore be a confounder, collider, and/or mediator in the relation between brain structure and ADHD, as depicted in the DAGs in **Figure 5** and **Supplementary Box 2**.

First, it could be argued that IQ is partly innate and precedes brain development and ADHD, making it a confounder (**Figure 5A**). Second, IQ may lie in the pathway between brain structure and ADHD and therefore act as a mediator (**Figure 5B**). It is conceivable that cognitive differences, as a consequence of subtle neurodevelopmental differences (25), could underlie ADHD. Adjusting for a mediator would lead to bias when estimating the total association between brain structure and ADHD (26). Third, brain structure may impact intelligence scores (25), and ADHD symptoms may affect IQ test performance (27) (**Figure 5C**). A variable that is independently caused by the outcome and the determinant is also known as a collider, and adjusting for it leads to (collider) bias. Here, we explored the impact of adjusting for IQ when examining the relation between brain morphology and ADHD (model 4).

**Figure 5.**
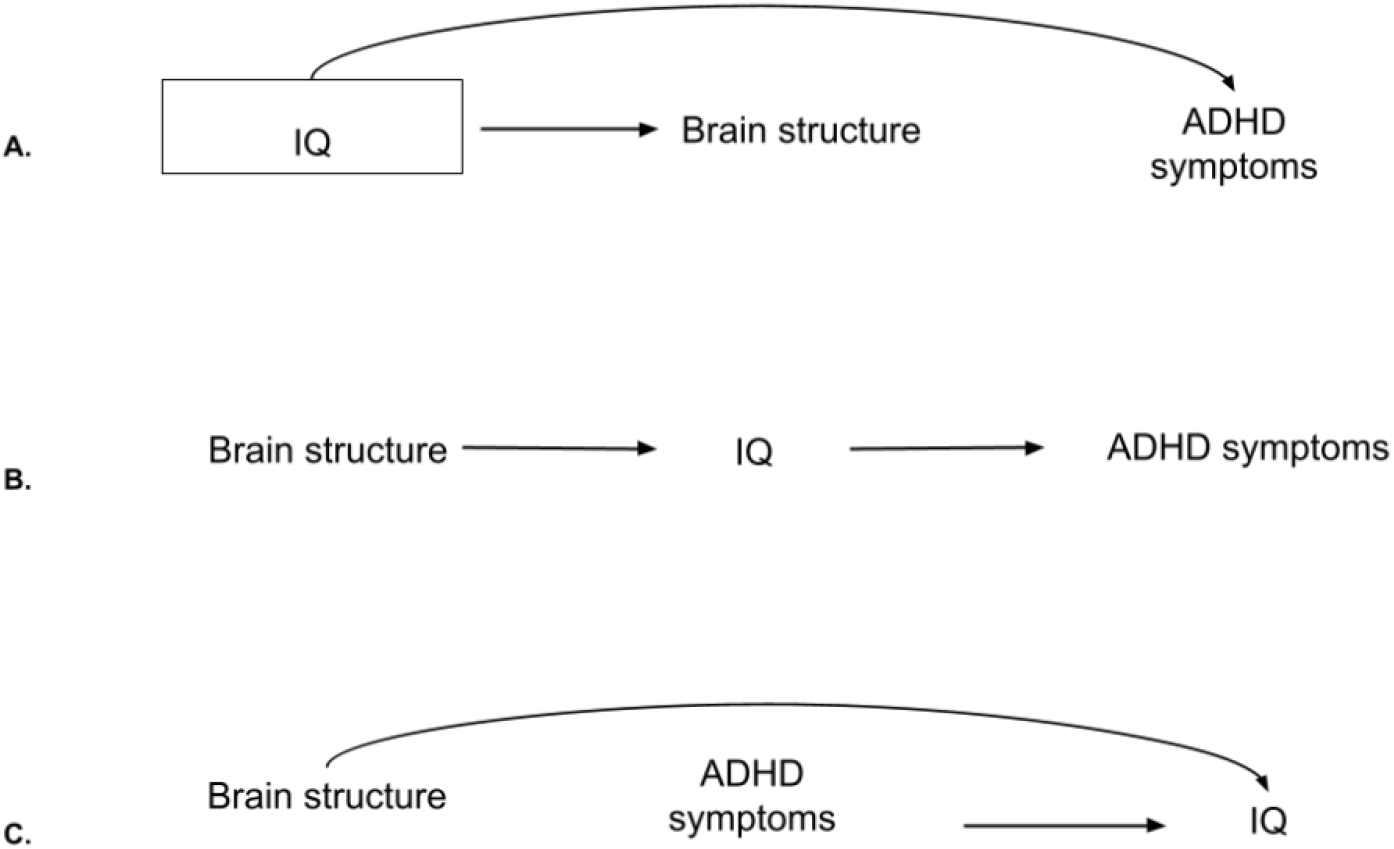
Directed acyclic graphs for IQ, brain structure, and ADHD symptoms. *Note*.**A)** DAG for IQ as a confounder. In this case, adjustments are needed as the backdoor path from brain structure to ADHD symptoms through IQ is open. By adjusting (box around IQ), the path gets closed. **B)** DAG for IQ as a mediator. Adjustments are not needed to estimate the total effect of brain structure on ADHD symptoms. **C)** DAG for IQ as a collider. The backdoor path through IQ is already closed. Adjustments would open the path and lead to collider bias.

### Adjustments for IQ led to further cluster reductions

After additionally adjusting for IQ, the spatial extent of the clusters associated with ADHD symptoms reduced further in both cohorts **(Figure 6**). For surface area, compared to model 3, clusters reduced from 682.6 cm^2^ to 525.7 cm^2^ (= - 23%) for the ABCD Study, and from 214.1 cm^2^ to 93.1 cm^2^ for the Generation R Study (= - 57%). For volume, compared to model 3, clusters reduced from 262.5 cm^2^ to 164.1 cm^2^ (= - 37%) in the ABCD Study and from 33.8 cm^2^ to 17.9 cm^2^ (= - 47%) in the Generation R Study.

**Figure 6.**
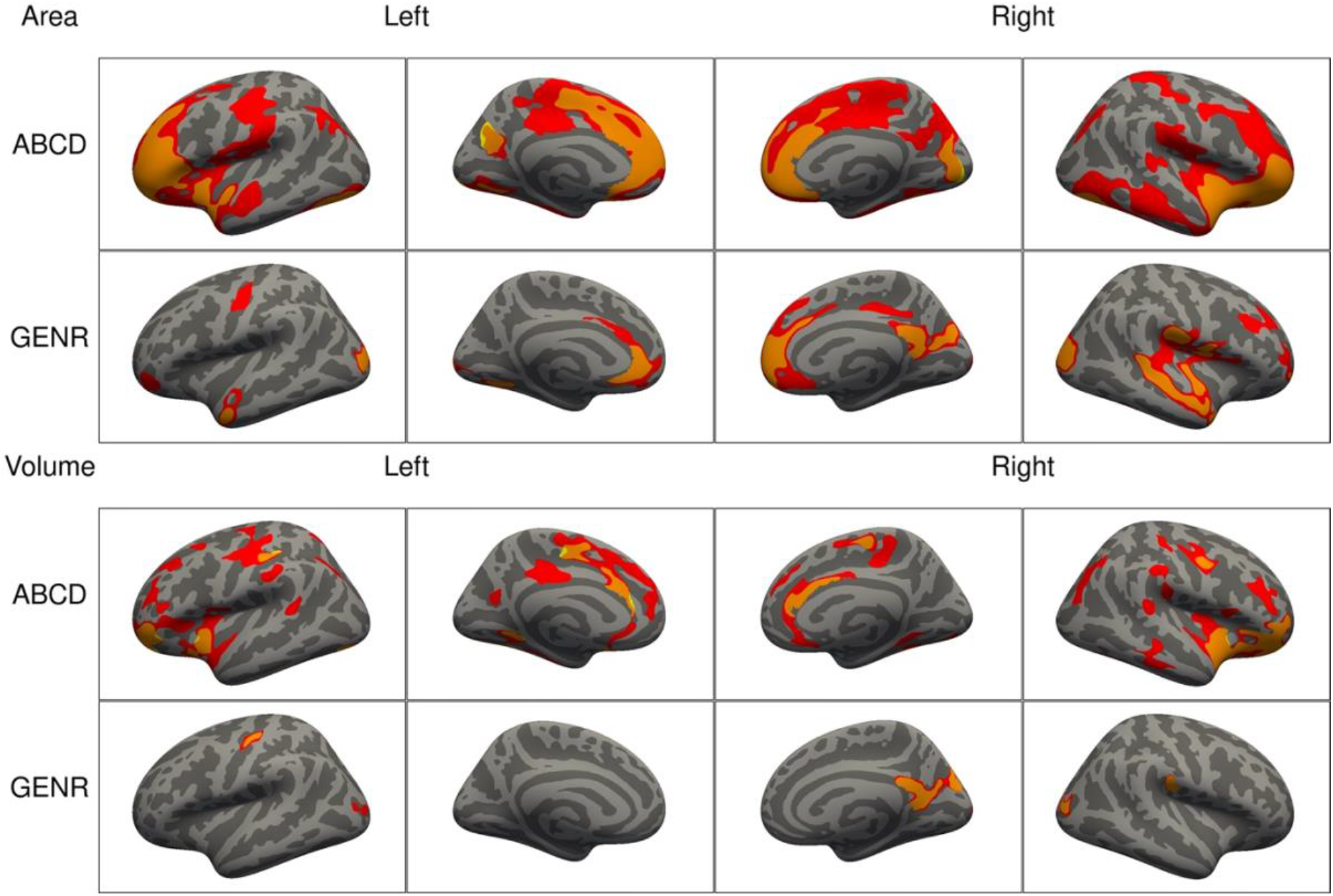
Significant clusters in the association of ADHD symptoms with cortical surface area (top) and volume (bottom) based on the ABCD and Generation R Studies, after additional adjustment for IQ. *Note*. Rows represent the results for the ABCD or Generation R Studies, and the columns represent the left and right hemispheres. The colors denote the different models, with red vertices being significant only in model 3, orange ones in both model 3 and after adjustment for IQ, and yellow ones only after adjusting for IQ.

For both cohorts, clusters of association for surface area in model 4 were located in the fusiform, inferior parietal, insula, lateral occipital, middle temporal, pericalcarine, pre- and post-central, precuneus, rostral middle, and superior frontal, superior parietal and temporal, and supramarginal cortices. For volume, the remaining clusters consistently identified across cohorts were the isthmus of the cingulate, lateral occipital, pre- and post-central, lingual, precuneus, superior parietal, and supramarginal cortices.

## Discussion

By leveraging two large population-based studies and adopting a literature- and DAG-informed approach to address confounding, we showed that *(i)* associations between brain structure and ADHD symptoms, which were initially widespread, reduced when adjusting for socioeconomic and maternal behavioral confounders, and that *(ii)* careful considerations are needed when including IQ due to its unclear relation with ADHD and brain morphology.

### Adjustments for confounders highlighted key regions of association, observed across two large cohorts

Widespread associations between surface area and volume with ADHD symptoms were initially identified, with higher symptoms relating to smaller brain structures, in line with previous research (4,28). After adjustments for potential confounders typically overlooked by previous literature (socioeconomic and maternal behavioral factors), approximately half of the associations remained, and considerable effect size changes were observed in both the ABCD and Generation R Studies. We observed similar patterns of cluster reductions for the relation of ADHD diagnosis with surface area and volume in the ABCD Study.

Regions that remained associated after adjustments and which were consistently identified across cohorts were the precuneus, isthmus of the cingulate, supramarginal, pre- and post-central, and inferior parietal cortices for both area and volume. Most of these regions (e.g., supramarginal) have been previously implicated in ADHD in clinical samples (29–31). However, many different brain areas have been detected in association with the disorder (11), which may have hampered prior meta-analytic efforts to identify consistent neuroanatomical correlates for ADHD.

Here, we discerned associated areas likely subject to confounding bias from areas robust to socioeconomic and maternal behavioral factors, and replicable across two large cohorts. Comparisons with prior findings should be made with caution due to differences in study design, samples (clinical vs. population-based), and analytical methods. Importantly, we highlighted the opportunity for future studies to include covariates that go beyond age and sex, can help refine associations, and can be readily collected. Future studies may want to consider other confounding factors, depending on their research question, design, and assumed causal relations.

### Adjustments for IQ are often unnecessary when examining the relation between brain structure and ADHD

Avoiding bias from adjusting for variables that are not confounders is as important as identifying sources of confounding. Adjusting for mediators or colliders of the ADHD-brain structure relation would induce bias. Here, when adjusting for IQ, which plays an unclear role in brain structure – ADHD associations, cluster sizes reduced considerably in both the ABCD and Generation R Studies. This could indicate that IQ is a confounder, in which case adjustments would be necessary, or that IQ is a mediator or collider, in which case adjustments must be avoided.

First, based on previous literature and this study, the association between ADHD and IQ is relatively weak (14) (*r*_ABCD_ = −0.11, *r*_GENR_ = −0.14), but this does not necessarily make it a weak confounder as the strength of confounding is due to a variable’s relation with the exposure *and* outcome. Second, if brain structure and ADHD symptoms both cause cognitive changes, adjusting for IQ could induce collider bias, although this is also dependent on when IQ is measured relative to the exposure and outcome (8). Third, if brain structure determines cognitive functioning, which in turn affects ADHD symptoms (mediation by IQ), adjustments would also induce bias (26).

Given these scenarios, we recommend moving away from routinely adjusting for IQ in ADHD neuroimaging studies, and we highlight the need to carefully consider the causal model for a specific research question to determine whether IQ may confound associations.

### Generalization to psychiatric neuroimaging studies

Our considerations on confounding likely generalize to the psychiatric neuroimaging field, as several confounders considered here (e.g., SES) also relate to brain function and other psychiatric disorders (32–34). Similarly, other psychiatric disorders are also characterized by complex relations with IQ (35).

Confounding control is paramount to studies examining determinants of a phenotype, like ADHD. However, even in these studies, one may be tempted to conduct correlational research with limited confounding adjustments, and then speculate about biological causal mechanisms (36,37). Rather, we suggest leveraging prior literature and expert knowledge to identify and adjust for key confounders. This can help eliminate the influence of alternative mechanisms (to the ones hypothesized) on the relation of interest (8). Charting the assumed (causal) structures to identify confounders can be done through the use of tools such as DAGs (8). Naturally, the plausibility of such assumptions should be evaluated. To facilitate the minimization of confounding bias in psychiatric neuroimaging, we propose a workflow in **Figure 7**.

**Figure 7.**
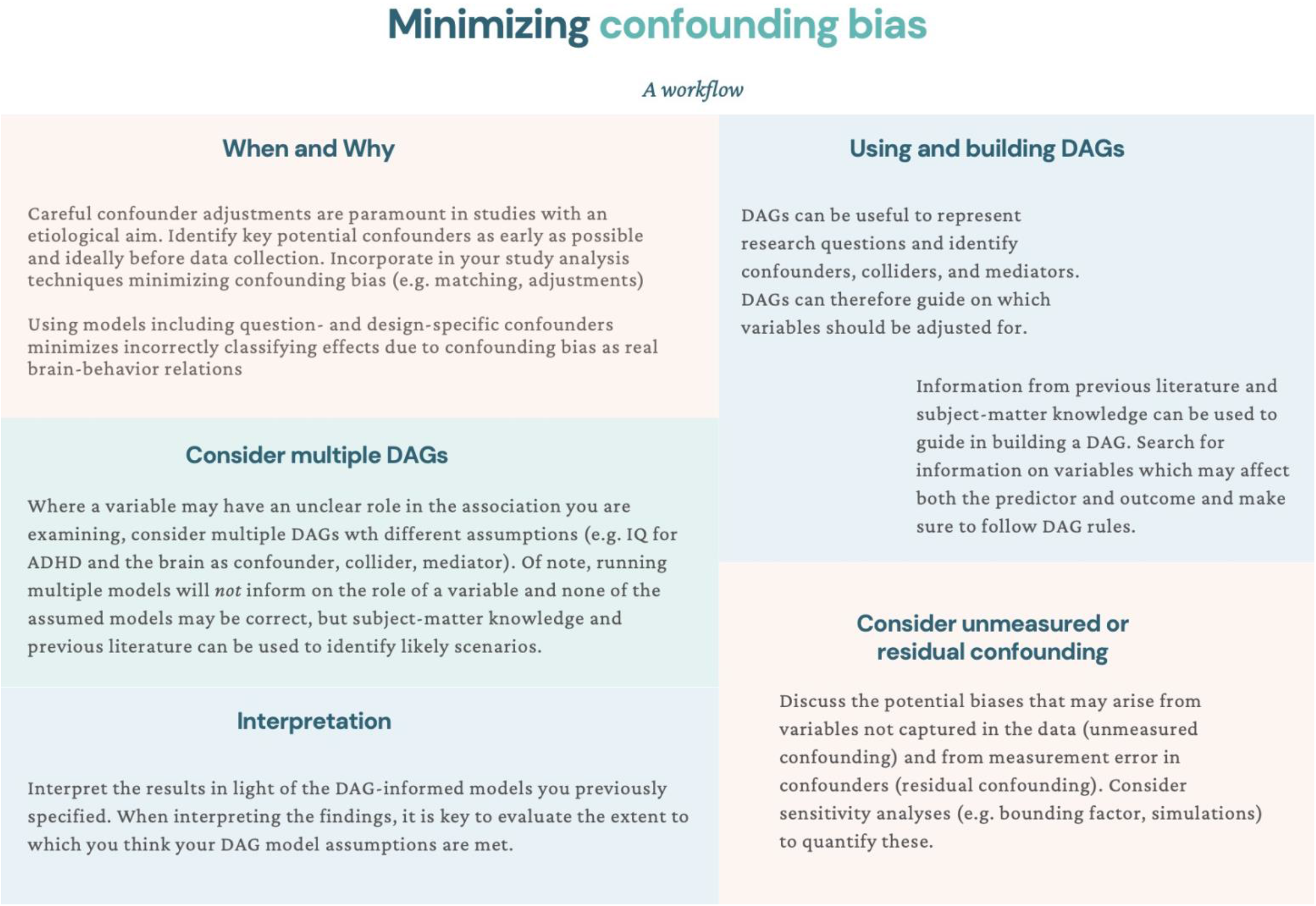
Suggestions for minimizing confounding bias: A workflow.

### Limitations of the present study and suggestions for future research

Despite leveraging two large samples with similar characteristics and assessments, this study presents several limitations. First, there is always potential for residual confounding through unmeasured confounders and misclassification of measured confounders. For example, given that genetic factors influence both ADHD and brain morphology and that there is a genetic correlation between ADHD risk and intracranial volume (38–40), certain genetic risk variants may be unmeasured confounders. However, we aimed to illustrate plausible confounding bias scenarios for ADHD and brain structure, and not to provide an exhaustive list of potential confounders, which may vary depending on the study population and research question. Future studies should also consider bias analyses to assess the impact that residual confounding may have on the study results (41). Bias analyses can help understand the minimum association strength an unmeasured confounder needs to have with the determinant and outcome to fully explain away the findings (42). Developments may be needed, however, for their adaptation to the neuroimaging field.

Second, due to our cross-sectional design, deliberately chosen to correspond to most neuroimaging studies, we must assume all confounders precede our determinant and outcome. This is a plausible assumption for the Generation R Study as, being a prospective birth cohort, we could ensure that the confounders here considered temporally preceded both ADHD and neuroanatomical assessments. However, this was not possible for the ABCD Study, which started sampling at child ages 9-10 years. Future research on the temporal relations between potential confounders, ADHD, and brain structure will aid the minimization of confounding bias when investigating the structural substrates of ADHD.

Third, neuroimaging parameters, such as head motion, were not considered here because they are discussed elsewhere (43), and generally fall under measurement error (information bias) rather than confounding bias. For instance, even when children with ADHD move more in the scanner compared to controls, determining lower image quality, head motion during scanning cannot cause changes in ADHD symptom levels. Future research aiming to increase precision in their estimates may, however, benefit from adjustments for this and other neuroimaging parameters.

Lastly, while we leveraged both symptom-level and diagnostic data for ADHD, this was done within population-based studies. Our results cannot, therefore, be generalized to a clinical population. Future research could examine the extent to which associations between brain structure and ADHD change after adjustments for likely confounders in clinical samples.

In conclusion, leveraging an empirical example from two large studies on neuroanatomy and ADHD symptoms, we highlighted the opportunity for future studies to consider further key confounders. These can be identified based on prior literature and causal diagrams as well as be readily collected, offering a feasible venue for future research. Adjusting for these potential confounders helped identify more refined cortical associations with ADHD symptoms, robust to the influence of demographic and socioeconomic factors, pregnancy exposures, and maternal psychopathology. We also evaluated the potential role of IQ, which could be a mediator, collider, and/or confounder. While adjusting for IQ led to reductions in associations, these would, however, likely not be attributable to reduced confounding bias. We discussed the generalizability of these considerations on confounding bias to psychiatric neuroimaging, and suggest a workflow that can be followed to minimize confounding bias in future studies.

## Methods and Materials

### Participants

We analyzed data from two independent population-based cohorts: The ABCD study and the Generation R Study. The ABCD study is conducted across 21 study sites in the US and recruited since 2015 children aged 9 to 10 at baseline (44). The Generation R Study is based in Rotterdam, the Netherlands, with data collection spanning from fetal life until early adulthood, and started in 2002 (16). Details of the sampling rationale, recruitment, methods, and procedures have been described elsewhere (16,44). Research protocols for the ABCD study were approved by the institutional review board of the University of California, San Diego, and the institutional review boards of the 21 data collection sites, while the design of the Generation R study was approved by the Medical Ethics Committee of the Erasmus MC. For both studies, written informed consent and assent from the primary caregiver or child were obtained.

In this cross-sectional study, we leveraged data from the baseline assessment of the ABCD study (release 2.0.1) and the 10-year assessment of the Generation R study. Both waves included behavioral and neuroimaging measures. We included children with data on ADHD symptoms and T_1_-weighted MRI images. Participants were excluded if *(i)* they had dental braces, *(ii)* incidental findings, *(iii)* their brain scans failed processing or quality assurance procedures, or *(iv)* they were twins or triplets. Of note, excluding children with dental braces is unlikely to determine selection bias by SES in either the ABCD or the Generation R study as the former cohort covered the costs of dental braces removal for all children who enrolled, while dental care is insured for all children in the Netherlands. Within the Generation R study, a small set of participants were additionally excluded because they had a different scan sequence. Finally, for each non-twin sibling set, one was randomly included to minimize shared method variance bias. Flowcharts for participant inclusion and exclusion are available in **Supplementary Figure 9**. The final samples consisted of 7,961 and 2,531 children from the ABCD and Generation R studies, respectively.

### Measures

#### ADHD symptoms

Children’s ADHD symptoms, reported by the primary caregiver, were measured with the CBCL (school-age version) (45), an inventory widely used for parent reports of children’s emotional and behavioral problems. The attention problem syndrome scale (20 items) measures inattention, hyperactivity, and impulsivity and has been previously shown to have clinical utility and to discriminate between ADHD cases and controls (46). Attention problems were analyzed on a discrete scale (range 0-19). For the ABCD study, we repeated the analysis using present ADHD diagnosis from a parent-reported and computerized version of the KSADS-5. This is a dimensional and categorical assessment used to diagnose current and past psychiatric disorders according to the Diagnostic and Statistical Manual of Mental Disorders (Fifth Edition) (47,48).

#### Image acquisition

T_1_-weighted data were obtained on multiple 3T scanners in the ABCD study (Siemens Prisma, General Electric (GE) 750 and Philips) and one scanner in the Generation R study (GE MR750w). Standard adult-sized coils were used for the ABCD study and an eight-channel receive-only head coil for the Generation R study. To acquire T_1_-weighted structural images, the ABCD study used an inversion prepared RF-spoiled gradient echo scan with prospective motion correction while the Generation R study used an inversion recovery fast spoiled gradient recalled sequence (GE option = BRAVO, TR = 8.77 ms, TE = 3.4 ms, TI = 600 ms, flip angle = 10°, matrix size = 220 × 220, field of view = 220 mm × 220 mm, slice thickness = 1 mm, number of slices = 230, ARC acceleration factor = 2). More details can be found elsewhere (15,49,50). Of note, in the ABCD study, a technical mistake occurred at one collection site, causing the hemisphere data to be flipped. This was fixed before processing.

#### Image processing

FreeSurfer (version 6.0.0) was used for image processing, which involved *(i)* removal of non-brain tissue, *(ii)* correction of voxel intensities for B_1_ field inhomogeneities, *(iii)* tissue segmentation, and *(iv)* cortical surface-based reconstruction. Cortical surface maps were smoothed with a full width of a half-maximum Gaussian kernel of 10 mm. Within the ABCD Study, quality assessment was based on the quality control and recommended inclusion criteria for structural data from the ABCD team (49). Within the Generation R Study, quality assurance was manually performed by visually inspecting all images by trained raters, as previously described in the literature (51). Poor quality reconstructions were excluded.

#### Covariate assessment

##### The ABCD study

All data were collected at baseline (child age 9 to 10-years). Age and sex were recorded at intake. Child race/ethnicity was reported by the primary caregiver and was categorized as White, Black, Hispanic, Asian, Other by the ABCD team. Household combined net income (<$50,000, >=$50,000 & < $100,000, >=$100,000) and highest maternal education (<high school, high school diploma/GED, some college, bachelor degree, postgraduate degree) were self-reported by the primary caregiver in the Parent Demographics Survey. Maternal age at childbirth was measured in the Developmental History Questionnaire. Tobacco and cannabis use during pregnancy were retrospectively reported by the mother (yes, no, I do not know) in the Developmental History Questionnaire. Caregiver psychopathology was obtained from the Total Problems Adult Self Report Syndrome Scale. The Wechsler Intelligence Scale for Children-6 Matrix Reasoning total scaled score was used as a proxy for IQ.

##### The Generation R study

Age and sex were measured based on medical records obtained at birth. Child ethnicity (western, non-western) was assessed based on the parents’ birth country, in line with the Statistics Netherlands bureau. Maternal age at childbirth was prospectively measured. Family income and highest maternal education were obtained through prospective self-reports by the mother and/or father at child age 5 years. Maternal education was coded into low (no/primary education), intermediate (secondary school, vocational training), and high (Bachelor’s degree/University). Household net monthly income was classified as low (< 2000 euros), middle (2,000-3,200 euros), and high (> 3,200 euros). Maternal postnatal psychopathology, measured at child age six months, was prospectively reported by the mother based on the Brief Symptom Inventory questionnaire global severity index. Mothers prospectively reported smoking (never used, used) and cannabis use during pregnancy (no use vs. use during pregnancy). Non-verbal child IQ was measured at child age five years, based on the Snijders-Oomen Niet-Verbale Intelligentie Test (52), a validated Dutch non-verbal intelligence test.

#### Covariate selection

Similar covariates were grouped into confounding sets to minimize the number of tested models while including relevant confounders. Factors included in model 1 related to demographic and study characteristics (age, sex, ethnicity, and study site). Age and sex were selected as these have been previously adjusted for in previous neuroimaging studies of ADHD (**Supplementary Table 1**). Ethnicity was used as a proxy for differential health risk exposure among people of different ethnic groups. The study site was incorporated to account for location and scanner differences in the ABCD study.

Further potential confounders were selected based on previous literature and with the aid of DAGs, as described in the *Results* section. In model 2, variables indicating socioeconomic factors were included (maternal education, household income, maternal age at childbirth). Household income and maternal education are generally considered to measure childhood SES in health research (53). Maternal age at childbirth can additionally inform on the SES of the child by capturing part of the variance unexplained by income and education (e.g., younger mothers facing higher occupational challenges, highly educated mothers delaying childbirth (54)). In model 3, maternal factors from the prenatal and postnatal period were grouped (tobacco and cannabis use during pregnancy and maternal psychopathology) to measure early life exposures which may impact a child’s brain and psychiatric development.

### Statistical Analyses

The R statistical software (version 3.4.3) was used for all analyses. Missing data on covariates were imputed with chained equations using the *mice* R package (55). Linear vertex-wise analyses were performed with the *QDECR* R package (56), with surface area/volume and ADHD symptoms as variables of interest. Thickness was not examined due to previously reported null findings in the Generation R study (4), and because it adds little information on top of surface area and volume. Correction for multiple testing was applied by using cluster-wise corrections based on Monte Carlo simulations with a cluster forming threshold of 0.001, which yields false-positive rates similar to full permutation testing (57). A Bonferroni correction was applied to adjust for analyzing both hemispheres separately (i.e., *p* < 0.025 cluster-wise).

Our analyses for aim 1 involved three vertex-wise linear regression models, which progressively expanded to adjust for additional confounding factors. The first model focused on demographic covariates, the second on socioeconomic ones, and the third on maternal behavioral variables related to psychopathology and pregnancy exposures. These models were run for ADHD symptoms (in both the ABCD and Generation R studies) and ADHD diagnosis (in the ABCD study only, as sensitivity analysis). One additional model, building upon model 3, was run to address aim 2, to illustrate the consequences of adjusting for IQ. Given that IQ and ADHD were weakly correlated (*r*_*ABCD*_ = −0.11, *r*_*GENR*_ = −0.14), multicollinearity was not expected.

## Data Availability

All datasets for this article are not automatically publicly available due to legal and informed consent restrictions. Reasonable requests to access the datasets should be directed to the Director of the Generation R Study, Vincent Jaddoe, in accordance with the local, national, and European Union regulations. Data for The ABCD Study is already open and available in the NIMH Data Archive (NDA) (https://nda.nih.gov/) to eligible researchers within NIH-verified institutions. Data can be accessed following a data request to the NIH data access committee (https://nda.nih.gov/), which should include information on the planned topic of study. The request is valid for one year. Data use should be in line with the NDA Data Use Certification. The code used for this study is publicly available at https://github.com/LorenzaDA/ADHD_brainmorphology_confounding

https://nda.nih.gov/general-query.html?q=query=featured-datasets:Adolescent%20Brain%20Cognitive%20Development%20Study%20(ABCD)

## ACKNOWLEDGEMENTS

The ABCD Study is supported by the National Institutes of Health and additional federal partners under award numbers U01DA041048, U01DA050989, U01DA051016, U01DA041022, U01DA051018, U01DA051037, U01DA050987, U01DA041174, U01DA041106, U01DA041117, U01DA041028, U01DA041134, U01DA050988, U01DA051039, U01DA041156, U01DA041025, U01DA041120, U01DA051038, U01DA041148, U01DA041093, U01DA041089, U24DA041123, U24DA041147. All supporters are mentioned at https://abcdstudy.org/federal-partners.html. Participating sites and study investigators are shown at https://abcdstudy.org/consortium_members/. While ABCD investigators designed, implemented the study, and/or provided data, they did not participate in this manuscript. This work reflects the authors’ views, and may not reflect those of the NIH or ABCD investigators. The Generation R Study is supported by Erasmus MC, Erasmus University Rotterdam, the Rotterdam Homecare Foundation, the Municipal Health Service Rotterdam area, the Stichting Trombosedienst & Artsenlaboratorium Rijnmond, the Netherlands Organization for Health Research and Development (ZonMw), and the Ministry of Health, Welfare and Sport. Neuroimaging data acquisition was funded by the European Community’s 7th Framework Program (FP7/2008-2013, 212652, Nutrimenthe). Netherlands Organization for Scientific Research (Exacte Wetenschappen) and SURFsara (Cartesius Compute Cluster, www.surfsara.nl) supported the Supercomputing resources. Authors are supported by an NWO-VICI grant (NWO-ZonMW: 016.VICI.170.200 to HT) for HT, LDA, SL, and the Sophia Foundation S18-20, and Erasmus University and Erasmus MC Fellowship for RLM. We thank the participants, general practitioners, hospitals, midwives, and pharmacies in Rotterdam who contributed to the study.

## DISCLOSURES

We declare no conflicts of interest.

## STROBE Flowchart

Flowcharts of participant inclusion and exclusion for ABCD (panel A) and Generation R (panel B).

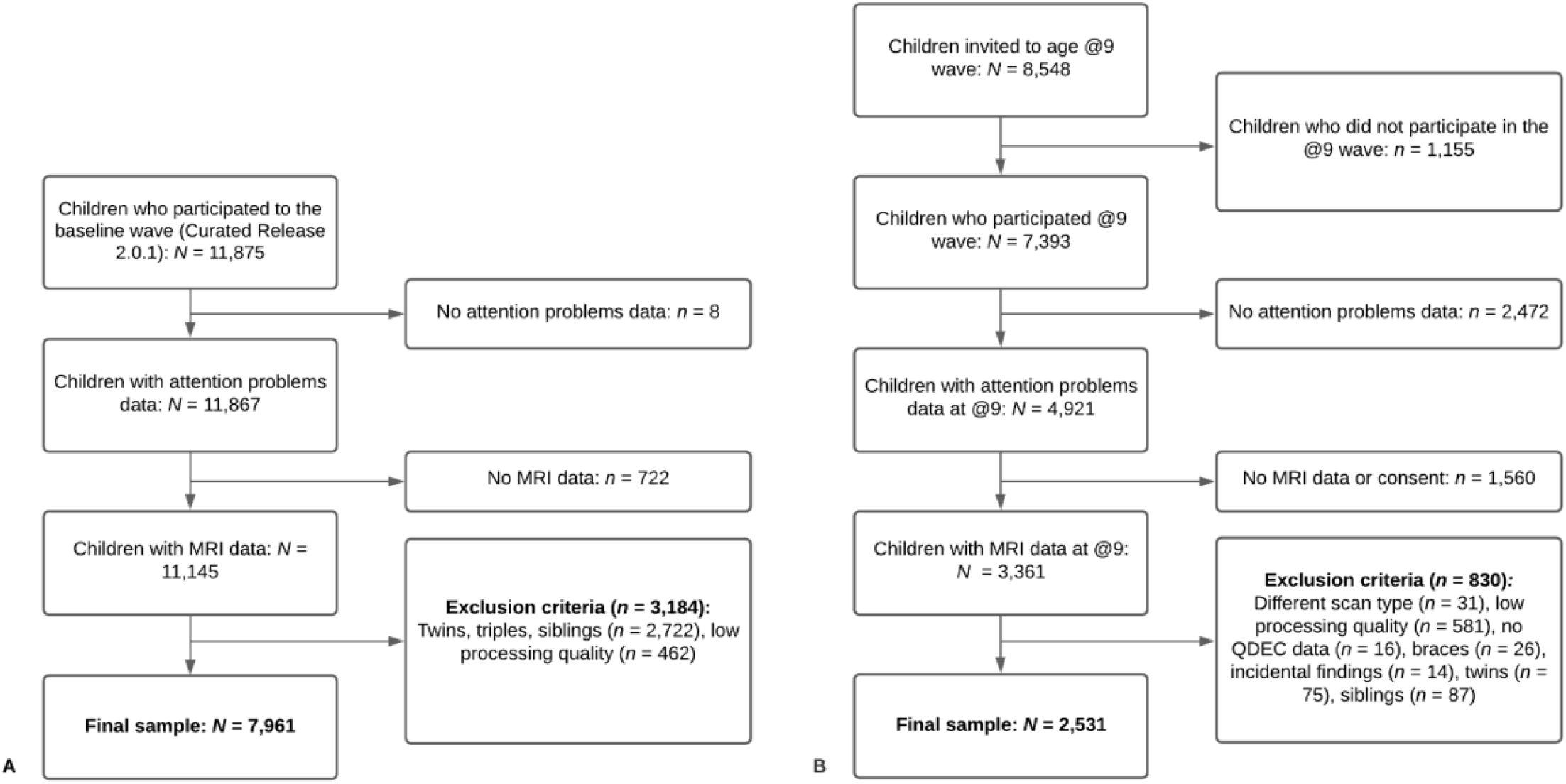

## Notes

### Competing Interest Statement

The authors have declared no competing interest.

### Author Declarations

Research protocols for the ABCD study were approved by the institutional review board of the University of California, San Diego, and the institutional review boards of the 21 data collection sites, while the design of the Generation R study was approved by the Medical Ethics Committee of the Erasmus MC.

